# The application of deep learning in lung cancerous lesion detection

**DOI:** 10.1101/2024.04.12.24305708

**Authors:** Phuong Thi Minh Chu, Tram Pham Bich Ha, Ngoc Minh Vu, Hoang Ha, Thu Minh Doan

## Abstract

**Background:** Characterized by rapid metastasis and a significant death rate, lung cancer presents a formidable challenge, which underscores the critical role of early detection in combating the disease. This study addresses the urgent need for early lung cancer detection using deep learning models applied to computed tomography (CT) images.

**Methods:** Our study introduced a unique non-cancer pneumonia dataset, a publicly available large-scale collection of high-quality pneumonia CT scans with detailed descriptions. We utilized this dataset to fine-tune nine pretrained models, including DenseNet121, MobileNetV2, InceptionV3, InceptionResNetV2, ResNet50, ResNet101, VGG16, VGG19, and Xception for the classification of lung cancer and pneumonia.

**Results:** ResNet50 demonstrated the highest accuracy and sensitivity (97.7% and 100%, respectively), while InceptionV3 excelled in precision (97.9%) and specificity (98.0%). The study also highlighted the contribution of the gradient-weighted class activation mapping (Grad-CAM) technique in examining the effectiveness of the model-training process via the visualization of features learned across different layers. Grad-CAM revealed that among the best-performed models, InceptionV3 successfully identified cancerous lesions in CT scans. Our findings demonstrated the potential of deep learning models in early lung cancer screening and improving the accuracy of the diagnosis procedure.

**Data availability:** The pneumonia CT scan dataset used in this study is extracted from peer-reviewed publications and can be accessed at https://github.com/ReiCHU31/CT-pneumonia-dataset

## 1. Introduction

Lung cancer remains the leading mortality globally, accounting for over 18% of all cancer-associated deaths in 2020 [1]. The symptoms of lung cancer are similar to common pulmonary diseases, and they often begin to stand out at advanced stages when the prognosis is less favorable [2]. Aside from clinical tests, non-invasive imaging techniques play an essential role in the screening and diagnosis of lung cancer. Patients are often prescribed chest X-rays, computed tomography (CT), positron emission tomography (PET), or magnetic resonance imaging (MRI) [3]. Among these, chest X-rays, also known as radiographs, are the most widely employed due to their simplicity and cost-effectiveness, yet they lack the high resolution required for early disease detection. MRIs, while offering exceptional image quality, are rarely used due to their high cost and stringent facility requirements, particularly posing challenges in resource-limited settings such as developing countries [4]. Meanwhile, CT scans provide significantly higher-resolution two-dimensional images that can reveal small pulmonary nodules, which are instrumental for early lung cancer diagnosis [5].

Despite the superiority in resolution, clinicians still face challenges when using CT scans to distinguish between lung cancer and other pulmonary diseases, particularly pneumonia. Also characterized by consolidation, pneumonia may mask the manifestations of lung carcinomas in CT scans, leading to a considerable delay of 3 to 5 months for accurately diagnosing lung cancer for patients with pneumonia symptoms [6–8]. Moreover, it typically requires physicians 9 to 12 years of training to effectively carry out the diagnostic process. In poor and developing countries, where professionals and healthcare resources are limited, it is crucial to develop automatic systems that alleviate these burdens to ensure timely and accurate diagnoses.

The development of data-driven models opens a promising area for systematically analyzing medical results, where artificial intelligence (AI) and deep learning (DL) techniques are utilized to detect various diseases based on imaging test results. Convolutional neural networks (CNN), a class of deep neural networks (DNN), are commonly employed, involving convolutional layers, pooling layers, and fully connected layers to extract features from input data and feed this information to a classifier [9]. Numerous methods based on CNNs for lung disease risk prediction and diagnosis have been introduced (Table 1). However, limited effort has been put into applying this technique in classifying lung cancer and other pulmonary diseases, including pneumonia and COVID-19, due to the lack of an official large-scale pneumonia dataset. A CADx-based attempt to handle the limited data issue used a dataset of 33,676 images of both CT and X-ray scans labeled as lung cancer, COVID-19, and normal lung, but only X-ray scans of pneumonia, and no official CT dataset of pneumonia was introduced [10]. Aside from the limited availability of data, another difficulty of utilizing cancerous CT scans as training data for AI models is the introduction of noise. Though most cancer patients develop community-acquired pneumonia during their illness, cancer datasets have not been tested to identify images containing both pneumonia and pulmonary cancer features [11, 12].

**Table 1.**
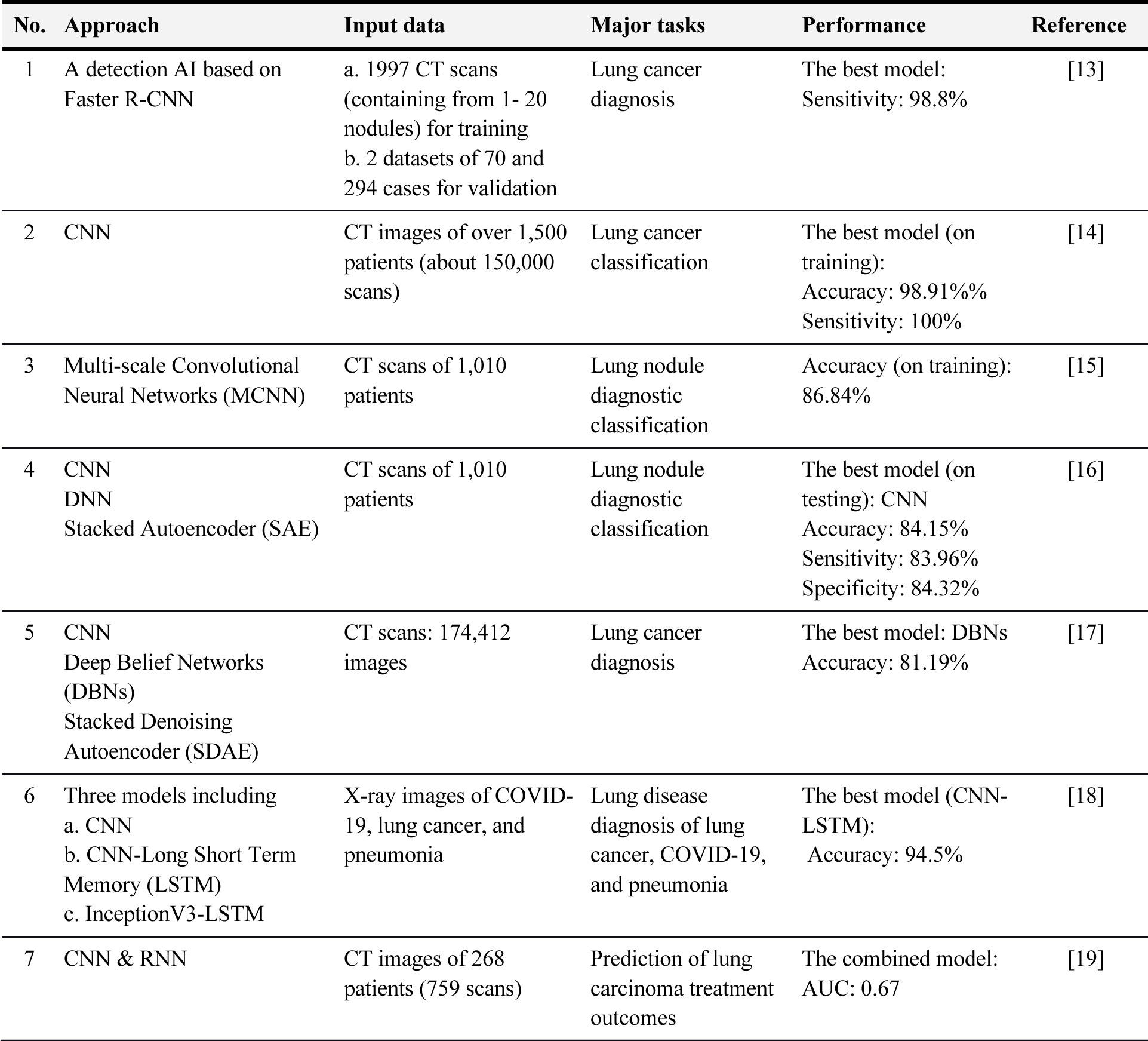
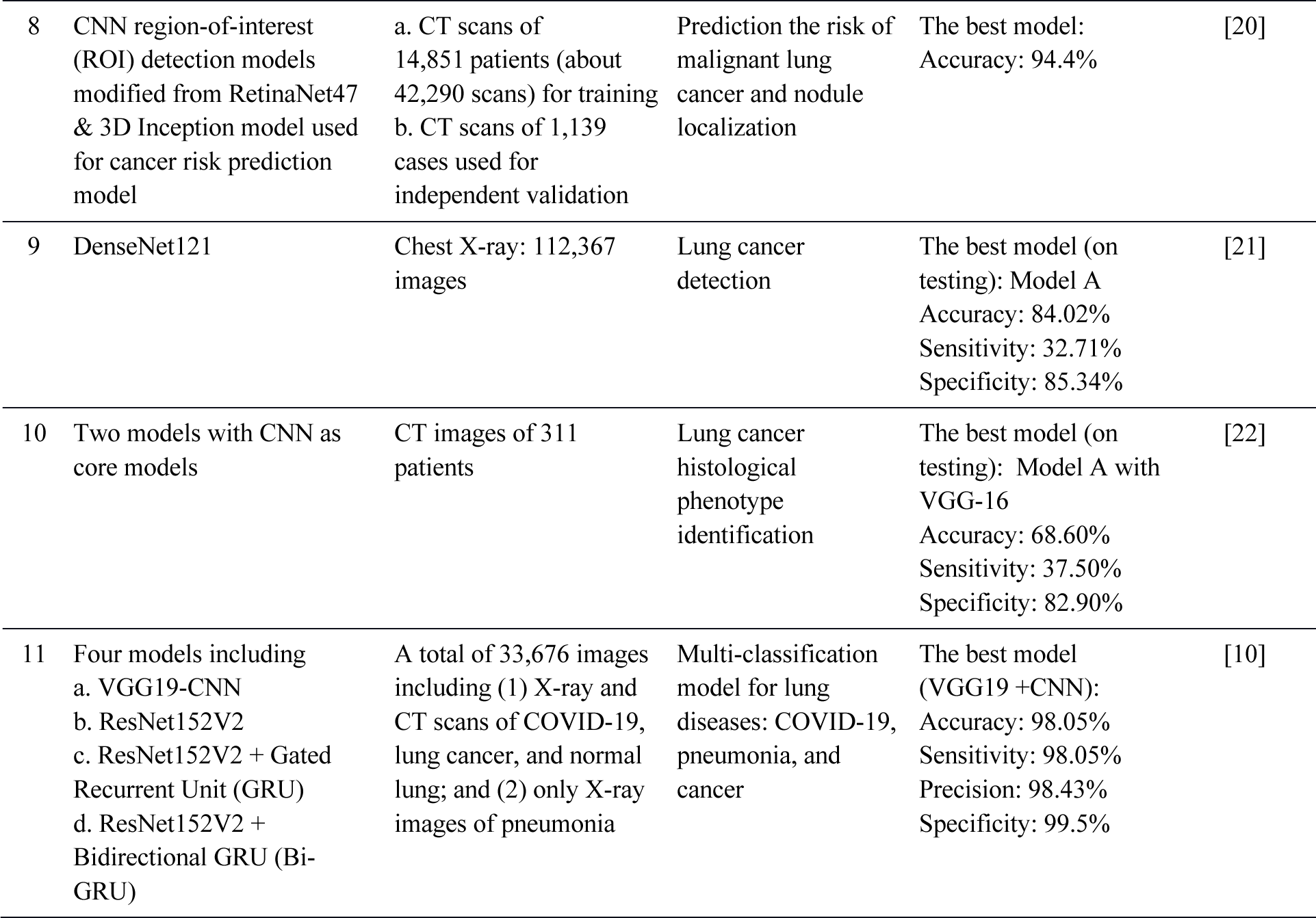
Current application of deep learning in lung cancer image diagnosis.

| No. | Approach | Input data | Major tasks | Performance | Reference |
| --- | --- | --- | --- | --- | --- |
| 1 | A detection AI based on Faster R-CNN | a. 1997 CT scans (containing from 1- 20 nodules) for training<br>b. 2 datasets of 70 and 294 cases for validation | Lung cancer diagnosis | The best model:<br>Sensitivity: 98.8% | [13] |
| 2 | CNN | CT images of over 1,500 patients (about 150,000 scans) | Lung cancer classification | The best model (on training):<br>Accuracy: 98.91%%<br>Sensitivity: 100% | [14] |
| 3 | Multi-scale Convolutional Neural Networks (MCNN) | CT scans of 1,010 patients | Lung nodule diagnostic classification | Accuracy (on training): 86.84% | [15] |
| 4 | CNN<br>DNN<br>Stacked Autoencoder (SAE) | CT scans of 1,010 patients | Lung nodule diagnostic classification | The best model (on testing): CNN<br>Accuracy: 84.15%<br>Sensitivity: 83.96%<br>Specificity: 84.32% | [16] |
| 5 | CNN<br>Deep Belief Networks (DBNs)<br>Stacked Denoising Autoencoder (SDAE) | CT scans: 174,412 images | Lung cancer diagnosis | The best model: DBNs<br>Accuracy: 81.19% | [17] |
| 6 | Three models including<br>a. CNN<br>b. CNN-Long Short Term Memory (LSTM)<br>c. InceptionV3-LSTM | X-ray images of COVID-19, lung cancer, and pneumonia | Lung disease diagnosis of lung cancer, COVID-19, and pneumonia | The best model (CNN-LSTM):<br>Accuracy: 94.5% | [18] |
| 7 | CNN & RNN | CT images of 268 patients (759 scans) | Prediction of lung carcinoma treatment outcomes | The combined model:<br>AUC: 0.67 | [19] |
| 8 | CNN region-of-interest (ROI) detection models modified from RetinaNet47 & 3D Inception model used for cancer risk prediction model | a. CT scans of 14,851 patients (about 42,290 scans) for training<br>b. CT scans of 1,139 cases used for independent validation | Prediction the risk of malignant lung cancer and nodule localization | The best model: Accuracy: 94.4% | [20] |
| 9 | DenseNet121 | Chest X-ray: 112,367 images | Lung cancer detection | The best model (on testing): Model A<br>Accuracy: 84.02%<br>Sensitivity: 32.71%<br>Specificity: 85.34% | [21] |
| 10 | Two models with CNN as core models | CT images of 311 patients | Lung cancer histological phenotype identification | The best model (on testing): Model A with VGG-16<br>Accuracy: 68.60%<br>Sensitivity: 37.50%<br>Specificity: 82.90% | [22] |
| 11 | Four models including<br>a. VGG19-CNN<br>b. ResNet152V2<br>c. ResNet152V2 + Gated Recurrent Unit (GRU)<br>d. ResNet152V2 + Bidirectional GRU (Bi-GRU) | A total of 33,676 images including (1) X-ray and CT scans of COVID-19, lung cancer, and normal lung; and (2) only X-ray images of pneumonia | Multi-classification model for lung diseases: COVID-19, pneumonia, and cancer | The best model (VGG19 + CNN):<br>Accuracy: 98.05%<br>Sensitivity: 98.05%<br>Precision: 98.43%<br>Specificity: 99.5% | [10] |

In this study, to explicitly distinguish patients from those with lung cancer and pneumonia, we collected chest CT scans from published scientific papers and open-access databases, then segmented the data into two categories: with lung cancer and pneumonia without lung cancer. The lung cancer data was extracted from The Cancer Imaging Archive (TCIA) [23, 24], while the pneumonia-only data was generated by collecting from peer-reviewed scientific publications. To our knowledge, our pneumonia CT scan dataset is the only published large-scale dataset compiled specifically for pneumonia detection. All CT scans were manually retrieved and image editing was performed to remove noise that may impede the classification process. Then, our team investigated whether lung cancer and pneumonia CT scans could be accurately differentiated by training nine deep-learning models. We adopted the fine-tuning approach, a transfer learning technique that takes advantage of models previously trained on a general domain, and then transfers the learned knowledge to another task of interest, allowing a high accuracy without the need for task-specific large-scale datasets or exhaustive training [25]. While a certain part of the pretrained weights was maintained, a custom classifier was applied on top of each deep neural architecture to achieve high accuracy and sensitivity. Furthermore, gradient-weighted class activation mapping (Grad-CAM) was implemented to create visual explanations for CNN models in the form of heatmaps. These heatmaps function as localization maps, highlighting regions within CT images that have a significant impact on the model’s decision-making process [26].

Our study successfully developed a new supplementary tool for the early detection and distinguishing lung tumors via (1) careful processing and noise removal in CT scans, (2) data augmentation for enlarging the dataset size, (3) optimizing and training CNN classification models and (4) using Grad-CAM for visual explanation of the models’ prediction process. Nine potential CNNs were trained and evaluated for automatic identification of whether a lung CT lesion includes cancerous regions with high accuracy, precision, and sensitivity.

## 2. Materials and Methods

### 2.1. Dataset building

Due to the lack of a publicly available database of pneumonia-only CT scans, a novel pneumonia dataset consisting of 1014 images was generated by curating CT scans from 192 peer-reviewed scientific literature and published databases. The CT images chosen are standard CT, contrast CT, and high-quality CT. The accepted types of pulmonary pneumonia are any infection due to bacteria, viruses, and fungi. The exclusion criteria are lung cancer, SARS-CoV-2 pneumonia, cryptogenic organizing pneumonia (COP), bronchiolitis obliterans organizing pneumonia (BOOP), and other lung damage that is not caused by bacteria, viruses, and fungi. All information, including the age and sex of patients, types of infection, and the source scientific articles, were recorded and stored in a metadata document. However, there is no information regarding the number of pneumonia patients those papers covered. Our complete dataset of pneumonia CT scans was established at https://github.com/ReiCHU31/CT-pneumonia-dataset.

For the cancerous dataset, a large-scale open-access library containing approximately 251,135 scans of 355 lung cancer patients has been employed [23, 24]. All lung cancer images in this study are collected from 101 random patients. The cancerous CT scans were evaluated by adept lung cancer radiologists to ensure validity. Clinical information was summarised in a metadata document containing patients’ ID, sex, age, weight, cancer stages, histopathological grading, and smoking history. Datasets’ means and medians are computed to understand the sample distribution. Our cancerous and pneumonia dataset consists of a total of 2028 CT scans, with an equal number of cases in each class.

### 2.2. Data processing and augmentation

All the collected CT scans were processed using Adobe Photoshop CS6 (64-bit) software to eliminate captions and annotations such as arrows or text symbols. These elements were not parts of the original images and were only useful in clinical examination, yet might introduce noise to the training data. Next, the edited CT images were labeled as one of the two classes: non-cancer (pneumonia) and cancer. The dataset was randomly split into a training set, validation set, and testing set. The training set consisted of 714 images of each class, while both the validation set and the testing set comprised 150 images of each class. To prevent overfitting and enhance model performance, data augmentation was performed in real time during the training process, in which CT images were subjected to random flipping, rotation, zooming, and distortion with varying degrees.

### 2.3. Model architecture and hyper-parameter tuning

The fine-tuning method was adopted to classify cancer versus non-cancer lung CT scans. To evaluate the effects of different model architectures for classification, we implemented 9 CNNs including DenseNet121, MobileNetV2, InceptionV3, InceptionResNetV2, ResNet50, ResNet101, VGG16, VGG19, and Xception. All models were pretrained on the ImageNet dataset [27]. Since the deepest layers of CNNs contain data-specific features while the first layers extract general features, in our experiments, 70% of the layers of each model were frozen to retain the pretrained weights, and only 30% last layers were set to be trainable. For each base architecture, the last fully connected layers were removed and replaced by our customized classifiers, which were designed and optimized specifically to achieve the highest possible accuracy and stability for each model. All classifiers consisted of an up-sampling layer (4×4), followed by batch normalization and fully connected layers (Fig. 1). During the optimization process, the number of layers and nodes were altered for each model to improve accuracy. Dropout layers with 0.3 to 0.5 probabilities were also applied for the majority of models to enhance their stability, except for VGG16 and VGG19 since they performed better and with lower final loss in our experiments.

**Figure 1.**
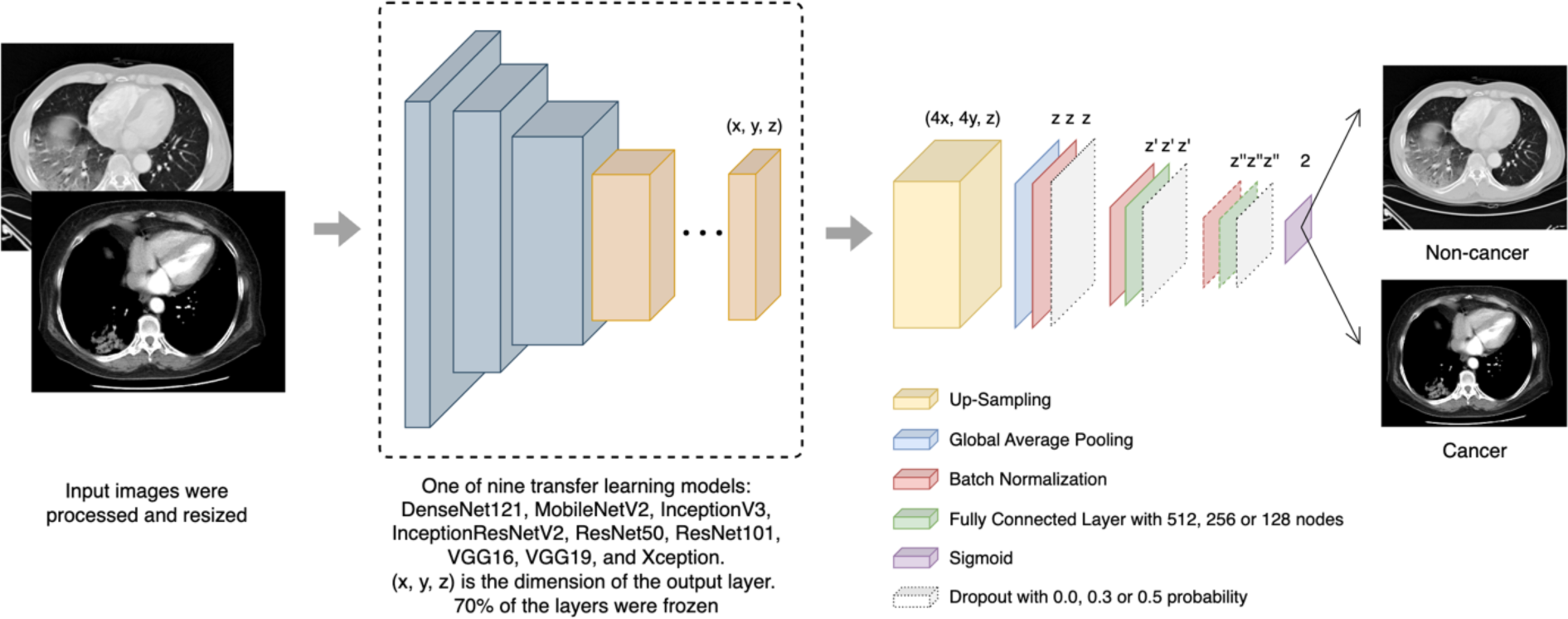
Overview of CNN architectures used in this study

Before training, all images in the dataset were resized to the default input size of each model according to the Keras API documentation [28], which was 224×224 for DenseNet121, ResNet50, ResNet101, 299×299 for InceptionV3, InceptionResNetV2, and Xception, which yielded desirable accuracy. However, for MobileNetV2, VGG16, and VGG19, we found the 320×320 image size delivered the best results and stability, therefore this size was used for these specific models. In addition, normalization of all images was performed according to each model’s requirements. Pixel values were either scaled to range from 0 to 1, from −1 to 1, or zero-centered without scaling.

All models were trained using the binary cross entropy loss function and Adam optimizer. Similar to the classifier optimization, the learning rate and epsilon values were adjusted for each model. Since our experiments follow the fine-tuning approach, low learning rates ranging from 5e-7 to 1e-3 were employed, and epsilon values were set from 0.001 to 0.1. All models were trained until converged using a batch size of 16 and epoch numbers ranging from 100 to 150.

The training process and optimization were performed using Google Colaboratory Pro’s NVIDIA Tesla T4s (50GB GPU, 32GB RAM) using Python 3.10 and Tensorflow Keras 2.15.

### 2.4. Model evaluation

All trained models were tested using a testing set containing 150 images of each class to evaluate the models’ performance on unseen data. The evaluation metrics including accuracy, precision, recall (i.e., sensitivity), specificity, and F1 score were calculated to assess model performance. The metrics were calculated as follows:

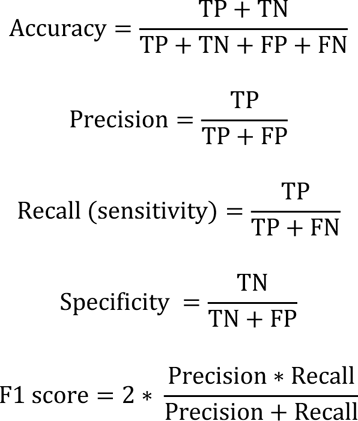

The true positives (TP), true negatives (TN), false positives (FP), and false negatives (FN) were derived directly from the confusion matrices that stored the classification results of the models when applied to the testing set. We consider true positives to be the data point correctly classified as cancer, and true negatives are those correctly predicted as pneumonia. In addition, we measured the area under the receiver operating characteristic curve (AUC), a metric used to assess the models’ performance across varying classification thresholds.

### 2.5. Gradient-weighted Class Activation Map (Grad-CAM)

After training the models according to the appropriate configuration, we applied Grad-CAM for all models using a sample of images from the dataset. While Grad-CAM can be applied to various layers within CNN models, the last convolutional layer is often preferred as it captures class-discriminative features that are crucial to a model’s decision on the input image [26]. This information was visualized with different color intensities in heatmaps for comparison between different models. Additionally, to evaluate the learned features in the CT scans across different learning stages, we averaged the gradients computed for both untrainable and trainable layers of each model. All Grad-CAM heatmaps were compared to assess whether meaningful insights could be derived from deep learning architectures, potentially informing faster and more accurate clinical diagnosis.

## 3. Results and discussion

### 3.1. Dataset evaluation

Two separate datasets were built from two primary sources (1) the pneumonia CT scans collected from scientific publications and (2) the Cancer Imaging Archive (TCIA) [23, 24]. This non-cancer pneumonia dataset is built differently from other datasets as it contains pneumonia scans only. Recent studies often used the input data that included COVID-19 CT images. To our knowledge, these COVID-19-included imaging data often lack detailed descriptions of clinical statuses such as the emergence of tumors or the signs of pneumonia caused by other pathogens rather than the SARS-CoV-2 viruses. We thoroughly dealt with many inadequacies of data to separate the two labels. The most significant challenge was the mixed input between cancer-only and cancer-with-pneumonia scans, which was extremely difficult to classify due to its inherently indistinguishable features reflected by CT images. After careful selection, two datasets including 2028 chest CT images, 1014 with a cancer label and 1014 with a non-cancer pneumonia-only label, were built. To date, our dataset is the only publicly accessible repository of pneumonia CT scans distinct from other pulmonary conditions.

The diversity of both datasets was also assessed. The lung cancer database includes all typical pulmonary cancer types, i.e., small cell lung cancer (SCLC) and non-small cell lung cancer (NSCLC, including adenocarcinoma, squamous cell carcinoma, and large cell carcinoma). The distribution of the data is similar to the actual disease rate in clinical settings. The 1014 lung cancer scans cover all stages of the TNM classification system, including four primary-tumor stages (T1, T2, T3, T4), four regional-lymph-nodes stages (N0, N1, N2, N3), and two metastasis-distance stages (M0, Mb) [29]. The non-cancer database is also an abundant collection of the infection causes with 32.56% bacterial, 48.64% viral, and 17.59% fungal pneumonia (Table S1). About 38.61% of cancer patients recorded in this dataset have a smoking habit. With the wide range of CT images, diverse pneumonia types, and without intended limitation in age, sex, lifestyle, or clinical status, the newly built pneumonia dataset allows the employed models to maximize the case-covering and the applicability in real-world situations.

### 3.2. Model evaluation

The novel dataset was used to train nine deep-learning models using the fine-tuning method to classify cancer and pneumonia from lung CT scans. 150 lung cancer CT scans and 150 pneumonia CT scans were used to evaluate all models. The performance of all models was assessed using accuracy, precision, recall, F1 score, specificity, and AUC, which were reported in Table 2. Each of these metrics provides insights into different aspects of the model’s capabilities. Our results revealed that all nine models achieved excellent performance across most metrics. ResNet50 stood out as the best model with an accuracy and F1 score of 97.7%, followed by VGG19 with 96.7% and 96.8% in these two metrics, respectively. Meanwhile, DenseNet121, despite its large number of layers and parameters, achieved lower accuracy (90.7%), precision (84.3%), F1 score (91.5%), and specificity (81.3%) compared to other models. Even though it demonstrated a comparable sensitivity value of 100%, the overall results indicated that DenseNet121 may be less ideal for our classification task.

**Figure 2.**
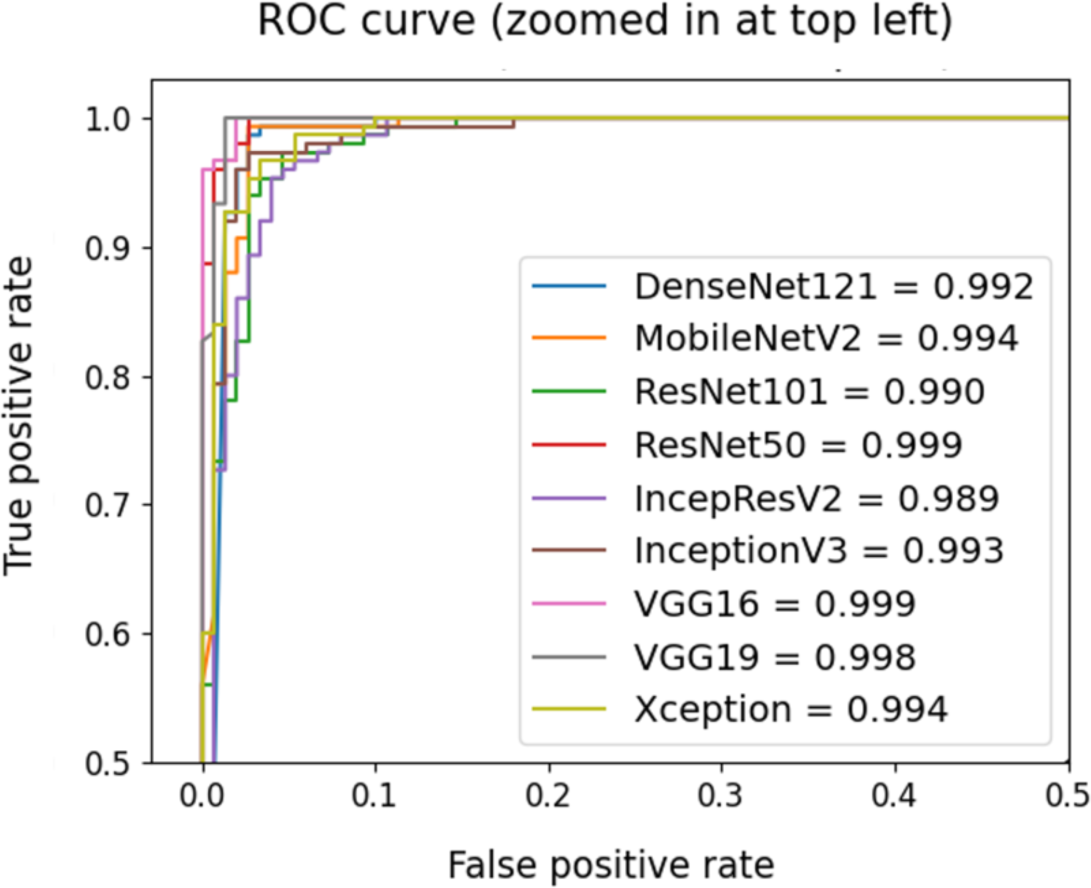
ROC curve and AUC values of all trained models

**Table 2.** Classification performance of all selected CNNs.

|  | Accuracy | Precision | Recall | F1 score | AUC | Specificity |
| --- | --- | --- | --- | --- | --- | --- |
| **DenseNet121** | 0.907 | 0.843 | **1.000** | 0.915 | 0.992 | 0.813 |
| **MobileNetV2** | 0.943 | 0.903 | 0.993 | 0.946 | 0.994 | 0.893 |
| **InceptionV3** | 0.967 | **0.979** | 0.953 | 0.966 | 0.993 | **0.980** |
| **InceptionResNetV2** | 0.937 | 0.888 | **1.000** | 0.940 | 0.989 | 0.873 |
| **ResNet50** | **0.977** | 0.955 | **1.000** | **0.977** | **0.999** | 0.953 |
| **ResNet101** | 0.963 | 0.954 | 0.973 | 0.964 | 0.990 | 0.953 |
| **VGG16** | 0.963 | 0.932 | **1.000** | 0.965 | **0.999** | 0.927 |
| **VGG19** | 0.967 | 0.938 | **1.000** | 0.968 | 0.998 | 0.933 |
| **Xception** | 0.963 | 0.948 | 0.980 | 0.964 | 0.994 | 0.947 |

Additionally, recall values, or sensitivity, remained exceptionally high for all models, with seven out of nine models achieving higher than 98%, indicating that most models were able to correctly identify all cancer cases in the testing set. This shows the effectiveness of our training approach since failing to detect cancer at an early stage might impede the diagnosis and treatment process in clinical settings. Models including InceptionV3, ResNet50, and ResNet101 exhibited high precision and specificity, with InceptionV3 being the top model with 97.9% and 98.0% in the two metrics. However, interestingly, InceptionV3 gained the lowest sensitivity of 95.3% out of all, indicating that though this model tended to accurately predict normal scans with minimum false positives, it might miss a slightly higher proportion of actual cancer-positive cases. A contrasting pattern was observed for VGG16 and VGG19. These models achieved perfect sensitivity, meaning they successfully identified all true cancer cases (with no false negatives). However, their specificity was much lower (92.7% for VGG16 and 93.3% for VGG19) compared to InceptionV3, due to classifying some pneumonia scans as cancer, leading to a higher number of false positives. This is a common trade-off between sensitivity and specificity. Ideally, high values of sensitivity and specificity suggest that the models were not only accurate in their classifications but also consistent in identifying both cancer-positive and negative cases correctly. Since our goal was to distinguish lung cancer cases from pneumonia cases to assist in timely diagnosis and treatment, models with higher sensitivity including ResNet50 and VGG19 might be more appropriate. Nonetheless, the F1 scores, a harmony of precision and recall, reported for the majority of models achieved more than 96%, indicating that our models are sufficient for the classification of cancer and non-cancer pneumonia CT scans.

We further confirm this observation by comparing the AUC values of all models (Figure 3). Rather than using a fixed decision threshold as the previously described metrics, AUC values correspond to the model’s ability to distinguish between positive and negative cases regardless of the chosen classification threshold [30]. Except for InceptionResNetV2, which gained 98.9% in AUC, all models achieved values higher than 99%. These findings collectively suggest that our deep learning models hold significant promise for aiding the classification of lung cancer and pneumonia from CT scans.

**Figure 3.**
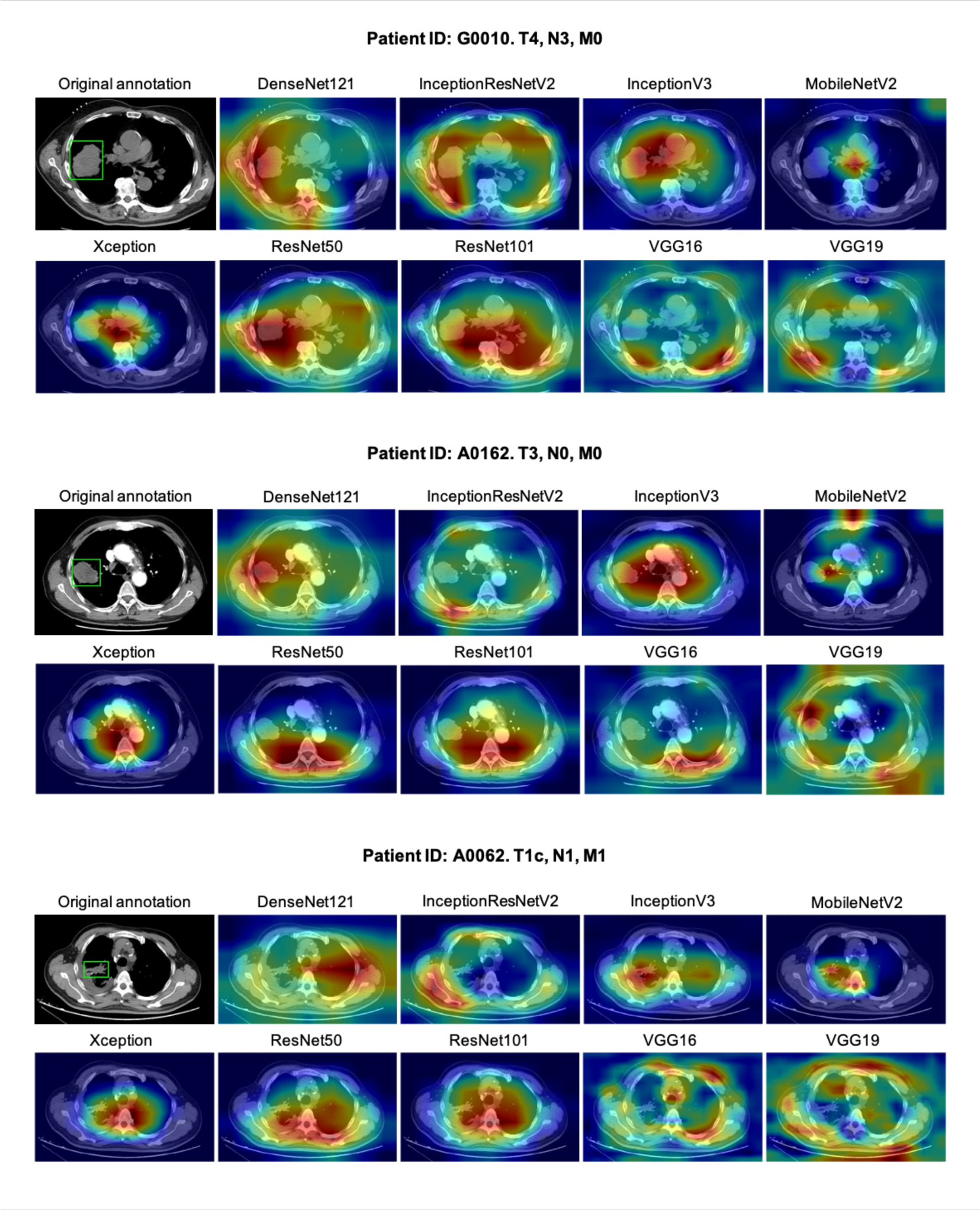
Grad-CAM visualization for cancer-positive CT scans using computed gradients from the last convolutional layer of each fine-tuned model. The features learned by each model are highlighted in a blue-yellow-red gradient, with red denoting the most focused regions in the image. In the original scans, each pulmonary tumor is denoted by a green bounding box. The coordinates of the bounding boxes were provided by the database’s annotation files.

We also took into account the number of parameters of each model to investigate whether this factor affected the training outcome. Our model architectures were optimized with custom layers, hence the difference in the number of parameters (Table S3). A large number of parameters allow deep neural networks to learn noise and small variations in the training data, which are known to cause overfitting, where the model fails to generalize for unseen data [31]. However, this was not the case for our models as evidenced by all evaluation metrics on the testing set, as well as the visualization of the training loss and validation loss throughout the training epochs, which demonstrated convergence in all models (Fig. S2). We also found no direct correlation between the number of parameters and overall accuracy. The best-performing models have moderate parameters, 24.1M for ResNet50, 20.1M for VGG19, and 22.3M for InceptionV3. While both ResNet50 and ResNet101 come from the same architecture family, with ResNet101 containing more layers and parameters, ResNet50 demonstrated a slight superiority in sensitivity and F1 score, indicating its potential for more accurate positive classifications. The performance of ResNet101 was comparable to Xception while containing more than twice the parameter counts. Notably, VGG19, having 5.3M parameters more than VGG16, showed a higher precision and F1 score.

In addition, the largest model is InceptionResNetV2 with 55.1M parameters, yet it achieved a modest precision of 88.8% and an accuracy of 93.7%. DenseNet121 and MobileNetV2 contain 7.3M and 2.6M parameters, respectively, and gained accuracies of 90.7% and 94.3%. Our results highlighted that specific model architecture and training strategies play a significant role in achieving high performance.

To assess the suitability of the models for clinical applications, where accuracy in disease classification is important as well as the ability to provide meaningful insights and interpretations of input images, we implemented the Grad-CAM technique. Grad-CAM helps identify which regions are activated when each model predicts a specific class, which are highlighted in the produced heatmap for each CT image. In clinical practice, CT scans are examined by experts to detect signs of lung cancer before moving toward further procedures. In cases of SCLC or NSCLC, the signs often include ground glass nodules, mass lesions (centrally or peripherally located), pleural effusion, bronchial narrowing, and wall thickening [32–34].

CT scans of three lung cancer patients with different tumor stages and lymph node involvement which were correctly classified as cancer by all nine models, were selected for Grad-CAM demonstration. We first examined the Grad-CAM heatmaps produced from the last convolutional layer of each model (Fig. 3). We considered the detection to be successful if the highlighted regions clearly indicate the tumor sites or partially cover more than 50% of the tumor. DenseNet121, despite yielding the lowest accuracy and precision, successfully detected the advanced pulmonary tumors while failing to address the stage T1c tumor. Models of the ResNet family showed a tendency to focus more on the lower lobes of the lung, with ResNet50 only correctly highlighting the tumor at the T4 stage. In contrast, MobileNetV2 was able to draw attention to the T1c cancerous lesion, yet only focused on the paratracheal nodes and the trachea rather than the advanced tumors in the remaining scans. A relatively similar tendency could be observed for Xception, which failed to highlight any signs of lung tumor.

Interestingly, InceptionV3 was the only model that consistently captured the pulmonary tumors in all three chosen CT images, with the highest coverage on the stage T1c scan. In the scans of advanced cancer patients, InceptionV3 was able to concentrate on the tumor, yet its highlighted regions also included the superior vena cava and the adjacent lymph nodes. Interpretation of these sites might require further inspections by expert radiologists, as N3 indicates the progressive spread of cancer to regional lymph nodes [35]. Surprisingly, well-performed models including VGG19 and VGG16 seemed to concentrate more on the periphery of the lung cavity and did not provide any direct clue regarding the position of the tumor.

Since each model was fine-tuned to produce the highest possible accuracy, the model’s ability to rely on indicative regions to classify cancer states may depend on the depth of each model. Deeper models with more layers are generally known for the ability to capture more abstract features, while the first layers usually focus on low-level information in the image (e.g., edges, shapes). We further assessed the features extracted throughout the model architectures using the CT obtained from the patient at the most advanced cancer stage (patient ID: G0010). The heatmaps were produced by leveraging gradients computed in different subsets of layers, including fine-tuned (trainable) layers, frozen layers, and all layers. Fig. S3 showed that the frozen layers effectively delineated anatomical structures such as the lung cavity, along with various internal features. Conversely, heatmaps generated from the fine-tuned layers tended to exhibit a more localized distribution of intensity, concentrating primarily on specific areas such as the tumor lesions or interior anatomical structures, indicating the model’s enhanced sensitivity.

The last convolutional layer is a common choice for Grad-CAM visualization, yet we saw that it does not represent the overall features extracted throughout the learning process of the model. Thus, we assessed Grad-CAM heatmaps produced by other layers from the fine-tuned architecture of ResNet50, VGG19, and InceptionV3. As shown in Fig. 4, each layer highlighted distinct regions in the CT scans with varying intensity. The tumor lesion was identified in layer *conv5_block3_2* of the ResNet50 model and was less pronounced in earlier layers. The attention of the VGG19 model was on various parts of the scans, including the tumor but did not specifically focus on this area. Meanwhile, for InceptionV3, the tumor and its adjacent sites were consistently emphasized across the selected layers, demonstrating its superiority in tumor detection compared to other models.

**Figure 4.**
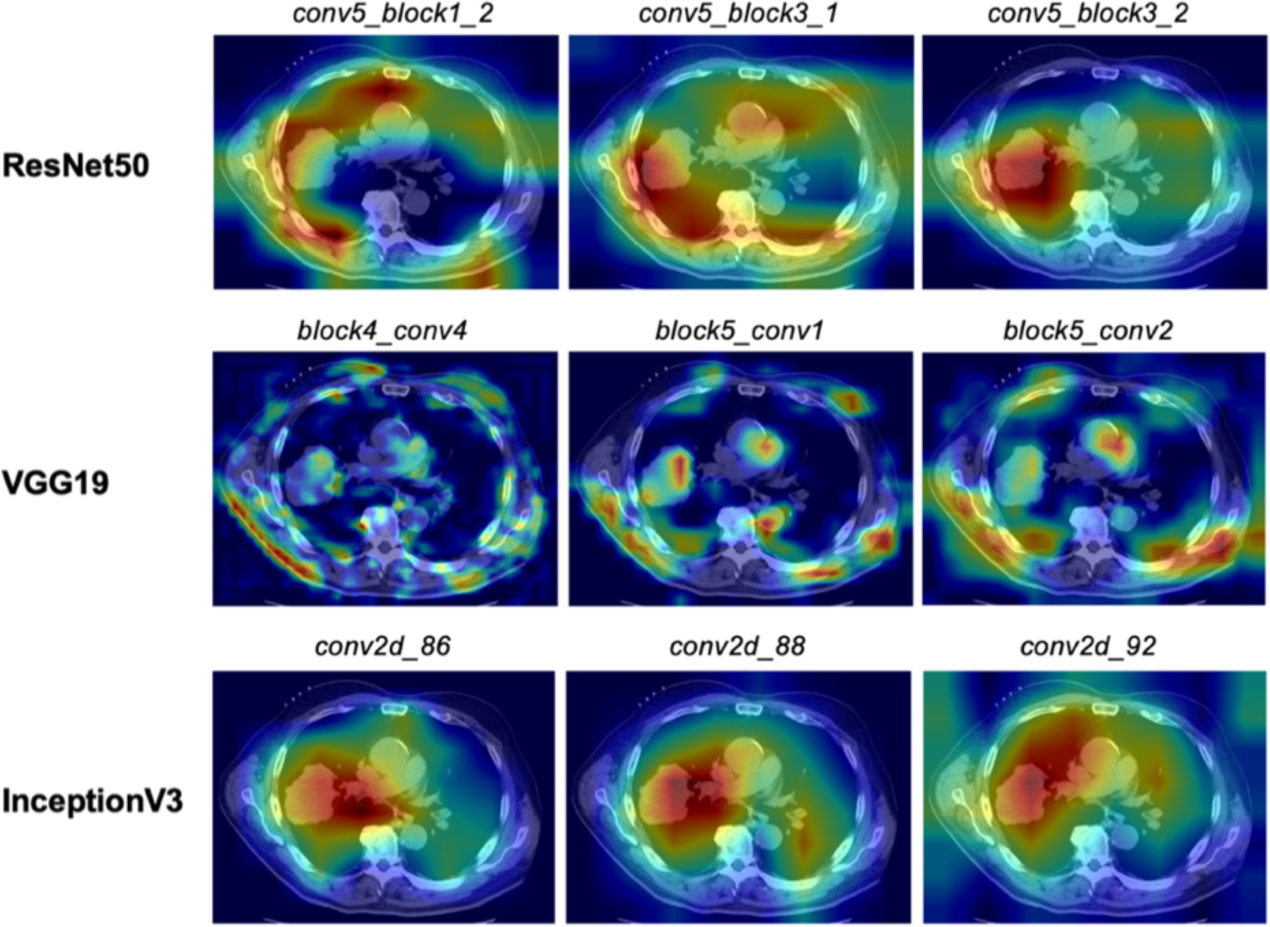
Grad-CAM heatmaps are derived from different layers of the three best models, namely ResNet50, VGG19, and InceptionV3.

Interpreting chest CT scans is an intricate process. Not only accurate and timely predictions of diseases but also the explainability of such decisions are needed for effective medical recommendations. Our results suggest that a combination of representations from various layers of classification models might be more beneficial in providing a more comprehensive understanding of the model’s disease predictions. Despite the excellent accuracy and sensitivity of our proposed models, including ResNet50 and InceptionV3, further calibration of model architectures might be essential to improve interpretability. In addition, our studies have not addressed the classification of lung cancer types (i.e., SCLC and NSCLC) versus pneumonia, which is crucial for early diagnosis. Thus, future investigations on this matter are crucial. Nonetheless, we believe incorporating the Grad-CAM heatmaps into the deep learning-based analysis process of lung CT scans would be useful in reducing diagnosis time and producing consistent classification results.

## 4. Conclusions

We introduced a curated dataset consisting of CT scans of pneumonia compiled from published scientific articles. Among other research on lung damage identification and classification, this work separated lung damage due to cancer from pneumonia by implementing nine deep-learning models using the fine-tuning approach. A total of nine CNNs were optimized and screened to determine the most appropriate architecture for the classification of lung cancer and pneumonia. All models achieved high accuracy, precision, sensitivity, and AUC, suggesting a promising computer-aided cancer detection and diagnosis. Particularly, InceptionV3 stood out with the highest precision and specificity and was also successful in detecting tumors in CT scans as indicated by Grad-CAM heatmaps. These results also recognized that CNN-based deep learning algorithms could be exploited to explicitly screen lung cancer and other diseases. Our newly established non-cancer pneumonia dataset is a valuable resource for future research in pulmonary CADe and CADx applications.

## Supporting information

Manuscript.pdf

## Data availability

The lung cancer CT image dataset is obtained from 101 random patients in a large-scale CT and PET/CT open-access library containing approximately 251,135 scans of 355 lung cancer patients [23, 24]. Meanwhile, the pneumonia dataset is a newly established library extracted from peer-reviewed scientific publications and is available at https://github.com/ReiCHU31/CT-pneumonia-dataset. It is imperative that this dataset be used solely for research purposes with responsibility. Ethical approval is not required. The results will be shared through various avenues, including peer-reviewed publications, conference presentations, and communication with other segments of healthcare and society.

## Code availability

The code for fine-tuning models and Grad-CAM implementation can be accessed at https://github.com/NgocVuMinh/Lung-Cancer-Pneumonia-Classification.git

## Acknowledgment

We thank Dr. Dat Huynh (Khoury College of Computer Sciences, Northeastern University) and Mr. Thanh Chi Lan Nguyen (Department of Electrical and Computer Engineering, Tufts University) for valuable discussions and reviewing of this work.

## Author contributions

**Phuong Thi Minh Chu:** Conceptualization, Data Curation, Methodology, Software, Formal Analysis, Validation, Writing – Original Draft Preparation. **Tram Pham Bich Ha:** Data Curation, Methodology, Formal Analysis, Software, Writing – Original Draft Preparation. **Ngoc Minh Vu:** Methodology, Software, Formal Analysis, Visualization, Writing – Review & Editing. **Hoang Ha:** Supervision, Writing – Review & Editing. **Thu Minh Doan:** Formal Analysis, Methodology, Validation, Writing – Original Draft Preparation.

## References

1. Sung H, Ferlay J, Siegel RL, Laversanne M, Soerjomataram I, Jemal A, Bray F (2021) Global Cancer Statistics 2020: GLOBOCAN Estimates of Incidence and Mortality Worldwide for 36 Cancers in 185 Countries. CA: A Cancer Journal for Clinicians 71:209–249

2. Feng J, Mu X, Ma L, Wang W (2020) Comorbidity Patterns of Older Lung Cancer Patients in Northeast China: An Association Rules Analysis Based on Electronic Medical Records. International Journal of Environmental Research and Public Health 17:9119

3. Dimastromatteo J, Charles EJ, Laubach VE (2018) Molecular imaging of pulmonary diseases. Respiratory Research 19:17

4. Ogbole GI, Adeyomoye AO, Badu-Peprah A, Mensah Y, Nzeh DA (2018) Survey of magnetic resonance imaging availability in West Africa. The Pan African Medical Journal. 10.11604/pamj.2018.30.240.14000

5. MacMahon H, Austin JHM, Gamsu G, Herold CJ, Jett JR, Naidich DP, Patz EF, Swensen SJ (2005) Guidelines for Management of Small Pulmonary Nodules Detected on CT Scans: A Statement from the Fleischner Society. Radiology 237:395–400

6. Søyseth V, Benth JŠ, Stavem K (2007) The association between hospitalisation for pneumonia and the diagnosis of lung cancer. Lung Cancer 57:152–158

7. Aquino SL, Chiles C, Halford P (1998) Distinction of consolidative bronchioloalveolar carcinoma from pneumonia: do CT criteria work? American Journal of Roentgenology 171:359–363

8. Marin AC, Prasad A, Patel V, Lwoodsky C, Hechter S, Imtiaz A, Patel P, Shah V, Appiah J, Cheriyath P (2023) Pulmonary Adenocarcinoma Mimicking Pneumonia in a Young Adult. Cureus. 10.7759/cureus.35267

9. O’Shea K, Nash R (2015) An Introduction to Convolutional Neural Networks.

10. Ibrahim DM, Elshennawy NM, Sarhan AM (2021) Deep-chest: Multi-classification deep learning model for diagnosing COVID-19, pneumonia, and lung cancer chest diseases. Computers in Biology and Medicine 132:104348

11. Islam KMM, Jiang X, Anggondowati T, Lin G, Ganti AK (2015) Comorbidity and Survival in Lung Cancer Patients. Cancer Epidemiology, Biomarkers & Prevention 24:1079–1085

12. Tanaka S, Inoue M, Yamaji T, Iwasaki M, Minami T, Tsugane S, Sawada N, Group the JS (2023) Increased risk of death from pneumonia among cancer survivors: A propensity score-matched cohort analysis. Cancer Medicine 12:6689–6699

13. Katase S, Ichinose A, Hayashi M, et al (2022) Development and performance evaluation of a deep learning lung nodule detection system. BMC Medical Imaging 22:203

14. Rossetto AM, Zhou W (2017) Deep Learning for Categorization of Lung Cancer CT Images. In: 2017 IEEE/ACM International Conference on Connected Health: Applications, Systems and Engineering Technologies (CHASE). pp 272–273

15. Shen W, Zhou M, Yang F, Yang C, Tian J (2015) Multi-scale Convolutional Neural Networks for Lung Nodule Classification. In: Ourselin S, Alexander DC, Westin C-F, Cardoso MJ (eds) Information Processing in Medical Imaging. Springer International Publishing, Cham, pp 588–599

16. Song Q, Zhao L, Luo X, Dou X (2017) Using Deep Learning for Classification of Lung Nodules on Computed Tomography Images. Journal of Healthcare Engineering 2017:e8314740

17. Sun W, Zheng B, Qian W (2016) Computer aided lung cancer diagnosis with deep learning algorithms. In: Medical Imaging 2016: Computer-Aided Diagnosis. SPIE, pp 241–248

18. ur Rehman A, Naseer A, Karim S, Tamoor M, Naz S (2023) Deep learning classifiers for computer-aided diagnosis of multiple lungs disease. Journal of X-Ray Science and Technology 31:1125–1143

19. Xu Y, Hosny A, Zeleznik R, Parmar C, Coroller T, Franco I, Mak RH, Aerts HJWL (2019) Deep Learning Predicts Lung Cancer Treatment Response from Serial Medical Imaging. Clinical Cancer Research 25:3266–3275

20. Ardila D, Kiraly AP, Bharadwaj S, et al (2019) End-to-end lung cancer screening with three-dimensional deep learning on low-dose chest computed tomography. Nat Med 25:954–961

21. Ausawalaithong W, Thirach A, Marukatat S, Wilaiprasitporn T (2018) Automatic Lung Cancer Prediction from Chest X-ray Images Using the Deep Learning Approach. In: 2018 11th Biomedical Engineering International Conference (BMEiCON). pp 1–5

22. Chaunzwa TL, Hosny A, Xu Y, Shafer A, Diao N, Lanuti M, Christiani DC, Mak RH, Aerts HJWL (2021) Deep learning classification of lung cancer histology using CT images. Sci Rep 11:5471

23. Clark K, Vendt B, Smith K, et al (2013) The Cancer Imaging Archive (TCIA): Maintaining and Operating a Public Information Repository. J Digit Imaging 26:1045–1057

24. Li P, Wang S, Li T, Lu J, HuangFu Y, Wang D (2020) A Large-Scale CT and PET/CT Dataset for Lung Cancer Diagnosis (Lung-PET-CT-Dx) [Data set]. 10.7937/TCIA.2020.NNC2-0461

25. Tammina S (2019) Transfer learning using VGG-16 with Deep Convolutional Neural Network for Classifying Images. IJSRP 9:p9420

26. Selvaraju RR, Cogswell M, Das A, Vedantam R, Parikh D, Batra D (2020) Grad-CAM: Visual Explanations from Deep Networks via Gradient-Based Localization. Int J Comput Vis 128:336–359

27. Deng J, Dong W, Socher R, Li L-J, Li K, Fei-Fei L (2009) ImageNet: A large-scale hierarchical image database. In: 2009 IEEE Conference on Computer Vision and Pattern Recognition. pp 248–255

28. Chollet F, others (2015) Keras.

29. Amin MB, Greene FL, Edge SB, Compton CC, Gershenwald JE, Brookland RK, Meyer L, Gress DM, Byrd DR, Winchester DP (2017) The Eighth Edition AJCC Cancer Staging Manual: Continuing to build a bridge from a population-based to a more “personalized” approach to cancer staging. CA: A Cancer Journal for Clinicians 67:93–99

30. Bradley AP (1997) The use of the area under the ROC curve in the evaluation of machine learning algorithms. Pattern Recognition 30:1145–1159

31. Salman S, Liu X (2019) Overfitting Mechanism and Avoidance in Deep Neural Networks. 10.48550/arXiv.1901.06566

32. Purandare NC, Rangarajan V (2015) Imaging of lung cancer: Implications on staging and management. Indian J Radiol Imaging 25:109–120

33. Kobayashi Y, Ambrogio C, Mitsudomi T (2018) Ground-glass nodules of the lung in never-smokers and smokers: clinical and genetic insights. Transl Lung Cancer Res 7:487–497

34. Lee D, Rho JY, Kang S, Yoo KJ, Choi HJ (2016) CT findings of small cell lung carcinoma. Medicine (Baltimore) 95:e5426

35. Rosen RD, Sapra A (2024) TNM Classification. StatPearls

