## Supplementary material for "The application of deep learning in lung cancerous lesion detection": Manuscript.pdf

**Table S1. Characteristics, including age, gender, and causes of pneumonia, of patients whose CT scans were used**

| | Lung cancer CT scans (n = 1014) | Non-cancer (Pneumonia) CT scans (n = 1014) |
|---|---|---|
| **Age (year)** | | |
| 0 – 44 | 7.89% | 40.24% |
| 45 – 54 | 18.73% | 13.02% |
| 55 – 64 | 29.59% | 9.17% |
| From 65 | 42.80% | 10.75% |
| Unknown | 0.99% | 26.82% |
| **Sex** | | |
| Female | 49.70% | 27.42% |
| Male | 50.30% | 45.56% |
| Unknown | 0.00% | 27.02% |
| **Causes of pneumonia** | | |
| Bacteria | N/A | 32.56% |
| Virus | N/A | 48.64% |
| Fungi | N/A | 17.59% |
| Mixed causes | N/A | 1.21% |

*N/A: not available

**Table S2. Model training parameters and classifier architectures**

| No. | Base model | Input size | Learning rate | Epsilon | Epochs | Classifier architecture |
|---|---|---|---|---|---|---|
| 1 | DenseNet121 | 224 | 6.00E-05 | 1.00E-02 | 120 | Up Sampling 2D (size=(4, 4))<br>Global Average Pooling 2D<br>Batch Normalization<br>Dropout (0.5)<br>Dense (256, ReLu)<br>Batch Normalization<br>Dropout (0.5)<br>Dense (2, Softmax) |
| 2 | MobileNetV2 | 224 | 1.00E-05 | 5.00E-03 | 120 | Up Sampling 2D (size=(4, 4))<br>Global Average Pooling 2D<br>Batch Normalization<br>Dropout (0.3)<br>Dense (256, ReLu)<br>Batch Normalization<br>Dropout (0.3)<br>Dense (2, Softmax) |
| 3 | InceptionV3 | 299 | 1.00E-05 | 1.00E-02 | 120 | Up Sampling 2D (size=(4, 4))<br>Global Average Pooling 2D<br>Batch Normalization<br>Dropout (0.3)<br>Dense (256, ReLu)<br>Batch Normalization<br>Dropout (0.3)<br>Dense (2, Softmax) |
| 4 | InceptionResNetV2 | 299 | 1.00E-04 | 7.50E-02 | 150 | Up Sampling 2D (size=(4, 4))<br>Global Average Pooling 2D<br>Batch Normalization<br>Dropout (0.5)<br>Dense (512, ReLu) |

| | | | | | | |
|---|---|---|---|---|---|---|
| | | | | | | Batch Normalization<br>Dropout (0.5)<br>Dense (2, Softmax) |
| 5 | ResNet50 | 224 | 1.00E-05 | 1.00E-02 | 100 | Up Sampling 2D (size=(4, 4))<br>Global Average Pooling 2D<br>Batch Normalization<br>Dropout (0.3)<br>Dense (256, ReLu)<br>Batch Normalization<br>Dropout (0.3)<br>Dense (128, ReLu)<br>Batch Normalization<br>Dropout (0.3)<br>Dense (2, Softmax) |
| 6 | ResNet101 | 224 | 1.00E-05 | 1.00E-02 | 120 | Up Sampling 2D (size=(4, 4))<br>Global Average Pooling 2D<br>Batch Normalization<br>Dropout (0.3)<br>Dense (512, ReLu)<br>Batch Normalization<br>Dropout (0.3)<br>Dense (256, ReLu)<br>Batch Normalization<br>Dropout (0.3)<br>Dense (2, Softmax) |
| 7 | VGG16 | 320 | 5E-06 | 1.00E-02 | 120 | Up Sampling 2D (size=(4, 4))<br>Global Average Pooling 2D<br>Batch Normalization<br>Dense (256, ReLu)<br>Batch Normalization<br>Dense (2, Softmax) |

| | | | | | | |
|---|---|---|---|---|---|---|
| 8 | VGG19 | 320 | 5E-06 | 5.00E-03 | 120 | Up Sampling 2D (size=(4, 4))<br>Global Average Pooling 2D<br>Batch Normalization<br>Dense (256, ReLu)<br>Batch Normalization<br>Dense (2, Softmax) |
| 9 | Xception | 299 | 1.00E-04 | 1.00E-01 | 120 | Up Sampling 2D (size=(4, 4))<br>Global Average Pooling 2D<br>Batch Normalization<br>Dense (256, ReLu)<br>Batch Normalization<br>Dropout (0.5)<br>Dense (128, ReLu)<br>Batch Normalization<br>Dropout (0.5)<br>Dense (2, Softmax) |

**Table S3. Number of trainable and non-trainable parameters of each fine-tuned model**

| | DenseNet121 | MobileNetV2 | InceptionV3 | InceptionResV2 | ResNet50 | ResNet101 | VGG16 | VGG19 | Xception |
|---|---|---|---|---|---|---|---|---|---|
| Total parameters | 7,305,538 | 2,592,578 | 22,337,058 | 55,132,898 | 24,155,138 | 43,850,370 | 14,849,602 | 20,159,298 | 21,428,906 |
| Trainable parameters | 3,154,882 | 2,075,842 | 14,005,890 | 24,335,074 | 17,776,002 | 23,993,602 | 11,932,418 | 14,292,226 | 11,118,346 |
| Nontrainable parameters | 4,150,656 | 516,736 | 8,331,168 | 30,797,824 | 6,379,136 | 19,856,768 | 2,917,184 | 5,867,072 | 10,310,560 |

### Figure S1: Curves of training accuracy and validation accuracy

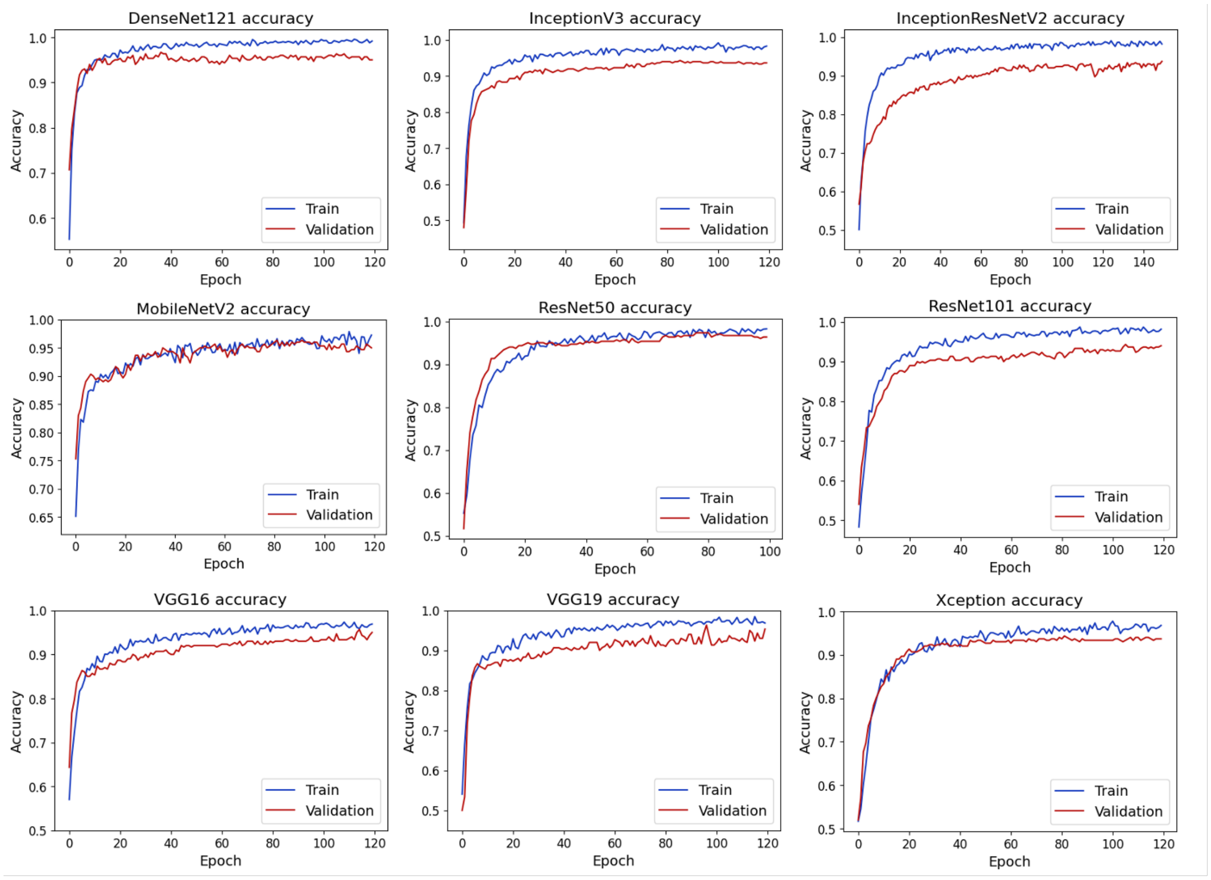

#### Figure S2: Curves of training loss and validation loss

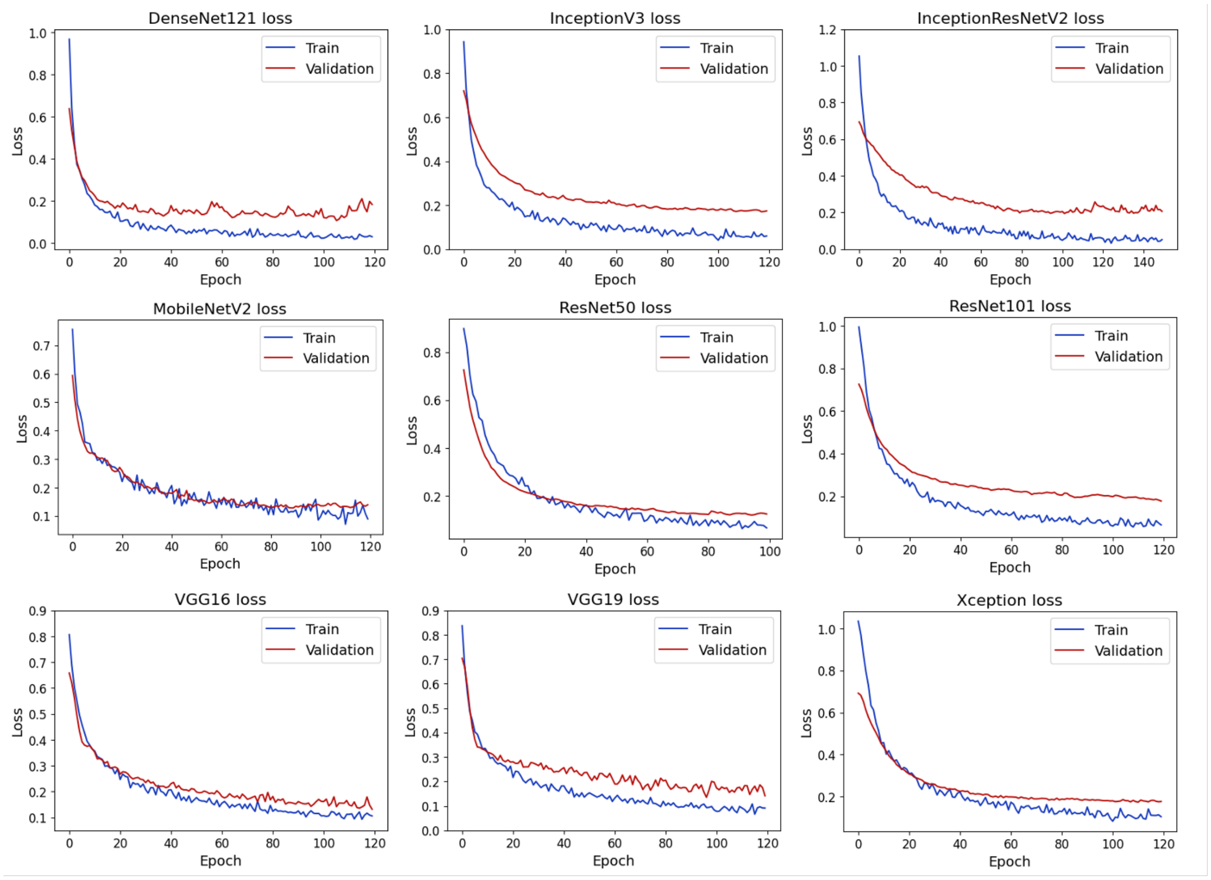

**Figure S3: Gradient-weighted Class Activation Map (Grad-CAM) for fine-tuned and pretrained layers of each model**

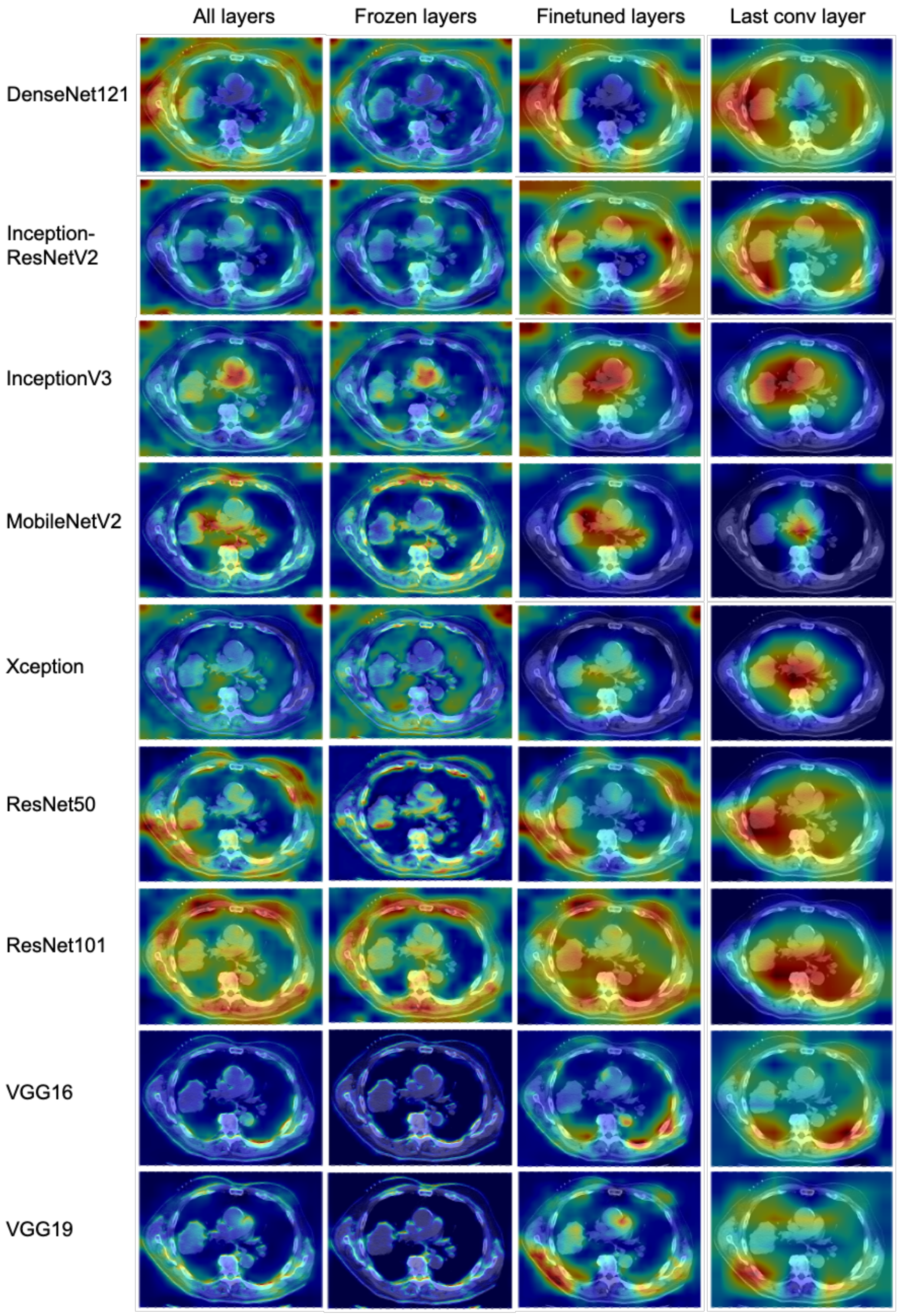
